# Spatiotemporal Clustering of Tuberculosis Notifications in Nepal During the COVID-19 Pandemic and Subsequent Recovery: A National Space-Time Scan Analysis, 2019–2024

**DOI:** 10.64898/2026.09.25.26364053

**Authors:** Nabin Bisht, Rupam Bhatt, Pratik Rijal, Ravi Kumar Gupta, Rekha Bisht, Harikishor Yadav, Samir Singh, Dilip Kumar Kalwar, Chiranjivi Adhikari

## Abstract

The COVID-19 pandemic substantially disrupted tuberculosis (TB) prevention, diagnosis, and treatment services worldwide. Despite increased TB notifications in Nepal during the post-pandemic recovery period, the geographic distribution and temporal clustering of notifications remain poorly understood. This study examined the spatial and temporal patterns of TB notifications across Nepal during the COVID-19 recovery period. We conducted a nationwide retrospective ecological analysis of district-level TB notifications across all 77 districts of Nepal from FY 2019/20 to FY 2023/24. Space-time, purely spatial, and purely temporal scan statistics were performed using a discrete Poisson model in SaTScan. Statistical significance was assessed using Monte Carlo simulation with 999 replications. A total of 172,155 TB cases were notified during the study period. TB notification rates increased by 51%, from 92 to 139 per 100,000 population between FY 2019/20 and FY 2023/24. Four statistically significant high-notification space-time clusters were identified (all p < 0.001). The primary cluster included Kathmandu, Lalitpur, Bhaktapur, Makwanpur, Bara, and Parsa districts during FY 2021/22–FY 2023/24 (RR = 1.60). Additional clusters were identified in western Nepal, the eastern Terai, and Rupandehi district. Purely temporal analysis showed a nationwide increase in notifications during FY 2022/23–FY 2023/24 (RR = 1.26, p = 0.001). TB notifications increased substantially during Nepal’s post-pandemic recovery period, with marked geographic and temporal heterogeneity. Integrating spatial and temporal surveillance may help identify areas requiring geographically targeted TB control strategies.

## INTRODUCTION

Tuberculosis (TB) remains one of the leading infectious causes of morbidity and mortality globally despite decades of coordinated international control efforts [1]. According to the World Health Organization (WHO), an estimated 10.7 million individuals developed TB and approximately 1.23 million died from the disease in 2024 [2]. South Asia bears a substantial proportion of the global TB burden, with several countries in the region classified among the WHO high-burden settings [3]. Persistent challenges in the region include delayed diagnosis, inequitable access to health services, multidrug-resistant TB, and socioeconomic vulnerability [3].

The COVID-19 pandemic severely disrupted TB control programs globally, with TB notifications declining by approximately 18% in 2020 and recovery remaining incomplete in subsequent years [2–4]. Lockdowns, diversion of healthcare resources, interruptions in diagnostic services, and reduced healthcare-seeking behaviour contributed to substantial declines in TB detection and treatment continuity [5,6]. Nepal is classified as a high-burden country for tuberculosis and multidrug-resistant TB [7].

National surveillance data indicate that TB notification rates had generally declined in the years preceding the COVID-19 pandemic, followed by substantial increases during the later study period [8]. However, national aggregate trends may obscure important geographic heterogeneity in TB notification patterns.

Spatial epidemiological approaches can identify geographic concentrations of infectious diseases and inform targeted public health interventions [9]. Space-time scan statistics provide a framework for detecting statistically significant disease clusters while accounting for spatial and temporal variation. Such methods have been widely applied to TB [10,11], dengue [12,13], COVID-19 [14] and other infectious diseases. Previous studies in Nepal have examined the spatial distribution of TB prevalence and notification rates, but available analyses have largely focused on cross-sectional or single-year patterns [15,16]. Evidence on the spatiotemporal dynamics of TB notifications in Nepal across the COVID-19 pandemic and subsequent recovery period remains limited.

Few studies have integrated space-time, purely spatial, and purely temporal analyses to characterize persistent geographic disparities alongside temporal changes in TB notifications. This study aimed to examine the spatiotemporal distribution of TB notifications across Nepal from FY 2019/20 to FY 2023/24 using national surveillance data and scan statistics. Specifically, we aimed to (1) identify statistically significant space-time clusters of elevated TB notification rates; (2) examine persistent geographic patterns of TB notifications; and (3) assess nationwide temporal trends in TB notifications across the study period, including the COVID-19 pandemic and subsequent recovery. Findings from this study may help inform geographically targeted TB control strategies and strengthen surveillance-based public health planning in Nepal.

## METHODS

### Study Design and setting

We conducted a retrospective ecological study of district-level tuberculosis (TB) notification data from all 77 districts of Nepal across five consecutive fiscal years, from FY 2019/20 through FY 2023/24 (July 16, 2019 to July 15, 2024). SaTScan was used to characterize spatial and temporal patterns of elevated TB notifications through three complementary analyses: retrospective space-time, purely spatial, and purely temporal scan statistics.

Nepal is a landlocked South Asian nation with a total population of approximately 29.1 million [17]. The country is administratively divided into seven provinces and 77 districts and encompasses three major ecological regions: Mountain, Hill, and Terai [18]. Districts were selected as the primary unit of analysis because they represent an operational level of Nepal’s National Tuberculosis Program (NTP) and are used for TB case notification, treatment monitoring, and drug supply management. District-level surveillance data were available across the entire study period.

For contextual interpretation of temporal patterns, the study period encompassed the COVID-19 disruption period and the subsequent recovery period. The COVID-19 disruption period was defined as FY 2019/20–FY 2021/22, encompassing the period during which COVID-19 and associated public-health measures substantially affected health-service delivery in Nepal. FY 2022/23–FY 2023/24 was considered the subsequent recovery period. These contextual periods were not used as predefined strata for the primary space-time scan analysis, which considered the entire FY 2019/20–FY 2023/24 study period.

### Data Sources

#### TB Surveillance Data

District-level TB notification data were obtained from the National Tuberculosis Control Centre (NTCC), Ministry of Health and Population, Government of Nepal [19]. The dataset included all notified TB cases recorded between July 16, 2019, and July 15, 2024, including bacteriologically confirmed pulmonary TB, clinically diagnosed pulmonary TB, and extrapulmonary TB cases reported by public and private healthcare facilities nationwide.

#### Population Data

District-level annual population estimates for FY 2019/20 to 2023/24 were obtained from the National Statistics Office (NSO) of Nepal [17]. These official population estimates were used as the population at risk and denominator for calculating annual TB notification rates.

#### Geographic Data

Administrative boundary shapefiles for Nepal’s 77 districts were obtained from the National Geoportal of Nepal [20]. Geographic coordinates were assigned using district centroids based on the World Geodetic System 1984 (WGS 1984). District identifiers (codes 101-709) were mapped to official district names based on the Government of Nepal’s administrative classification system (Appendix Table 1).

#### Outcome Variable

For descriptive analysis, the annual district-level TB case notification rate (CNR) was calculated as the number of notified TB cases per 100,000 population:

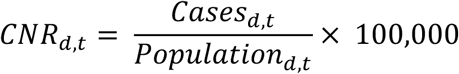

Where, d denotes district and t denotes fiscal year (FY 2019/20 to FY 2023/24). CNR was calculated for each district-year combination, producing 385 district-year records (77 districts × 5 fiscal years). For the scan-statistical analyses, notified TB cases were specified as the count outcome, with the corresponding population used as the population at risk.

#### Temporal Coding

Nepal’s fiscal year begins on July 16 (the first day of Shrawan in the Nepali Bikram Sambat calendar) and ends on July 15 of the following calendar year. For compatibility with SaTScan, which requires sequential positive integer values for temporal coding, the five fiscal years were coded sequentially from Period 1 to Period 5: Period 1 = FY 2019/20, Period 2 = FY 2020/21, Period 3 = FY 2021/22, Period 4 = FY 2022/23, and Period 5 = FY 2023/24 (Appendix Table 3).

### Statistical Analysis

#### Space-Time Scan Statistics

Retrospective space–time cluster analysis was performed using SaTScan version 10.3.2 [9,21]. A discrete Poisson model was specified because TB notifications were recorded as count data and population denominators were available for all districts. The space-time scan statistic uses a cylindrical scanning window that moves across geographic locations and time periods, with the circular base representing the spatial extent and the height representing the temporal duration of a potential cluster. For each scanning window, the observed number of TB notifications was compared with the expected number based on the population at risk in the corresponding districts and periods. A log-likelihood ratio (LLR) was calculated for each scanning window, and the window with the highest LLR was identified as the most likely cluster. Statistical significance was evaluated using 999 Monte Carlo replications. The relative risk (RR) quantified the relative rate of TB notification within a detected cluster compared with the area outside the cluster. An RR >1 indicated elevated notification risk within the cluster compared with the area outside the cluster.

#### Scan Parameter Specification

Scan parameters were defined a priori based on the epidemiology of tuberculosis (TB) in Nepal and recommendations from previous TB cluster studies [13]. The maximum spatial cluster size was set at 20% of the population at risk to reduce the likelihood of identifying very large clusters that could encompass geographically and epidemiologically heterogeneous areas [22, 23]. Sensitivity analyses using alternative spatial and temporal windows were conducted to assess the robustness of cluster detection. The maximum temporal cluster size was set at 60% of the study period (3 years) to allow detection of sustained multi-year clusters while avoiding clusters spanning most of the observation period. The minimum temporal cluster size was set at one year, corresponding to the annual reporting resolution of the dataset. A minimum of two cases was specified for cluster identification to reduce the likelihood of clusters based on very small numbers of notifications.

For primary reporting, clusters with an RR ≥1.5 were emphasized as having substantially elevated notification rates. This threshold was used as an epidemiological reporting criterion and was distinct from statistical significance, which was assessed using Monte Carlo simulation. Clusters with lower RR values were examined in sensitivity analyses. Circular spatial scanning windows were used because no specific directional pattern of geographic clustering was prespecified. Statistical significance was assessed using 999 Monte Carlo replications with an alpha level of 0.05.

### Complementary Analyses

Three complementary analyses were conducted. The space-time analysis identified geographic areas and periods with significantly elevated TB notification rates. The purely spatial analysis identified persistent geographic concentrations of elevated TB notification rates aggregated across the full study period. The purely temporal analysis assessed nationwide temporal variation in TB notifications independent of geographic location.

#### Sensitivity Analysis

Sensitivity analyses were conducted to assess the robustness of space-time cluster detection to alternative scanning-window specifications. The maximum spatial cluster size was varied between 20%, 30%, and 50% of the population at risk, while the maximum temporal cluster size was varied between 50% and 60% of the study period. The primary analysis used a 20% spatial window and a 60% temporal window, with clusters having RR ≥1.5 emphasized for primary reporting. Alternative specifications included 20% spatial 50% temporal, 30% spatial–60% temporal, and 50% spatial 60% temporal windows using the same RR ≥1.5 criterion. An 50% spatial 50% temporal analysis without an RR reporting restriction was performed to examine the broader spatial and temporal pattern of detected clusters. Additionally, a sensitivity analysis restricting the study period to FY 2021/22-FY 2023/24 (post-pandemic years 3–5 only) was conducted to assess whether identified clusters were detectable using post-pandemic data alone, independent of the pandemic disruption period. Robustness was assessed by comparing cluster location, temporal period, geographic extent, relative risk, and statistical significance across specifications [24]. Results from alternative parameter specifications are presented in Appendix Table 2.

#### Ethical Considerations

The Institutional Review Committee of Pokhara University, Nepal, has provided an exemption from ethical approval under reference number PU-IRC-36/2082/83.

## RESULTS

### Epidemic Overview

Between FY 2019/20 and FY 2023/24, 172,155 TB cases were notified across Nepal’s 77 districts. The national TB notification rate increased from 92 to 139 per 100,000 population, representing a 51% increase (Fig 1). Males consistently accounted for 61–63% of notifications. The proportion of bacteriologically confirmed pulmonary TB increased from 55.0% to 57.5%. The proportion of notified cases aged ≥65 years increased from 17.1% to 22.8%, while notifications among children aged 0–4 years increased from 651 to 2,007 (Table 1).

**Fig 1.**
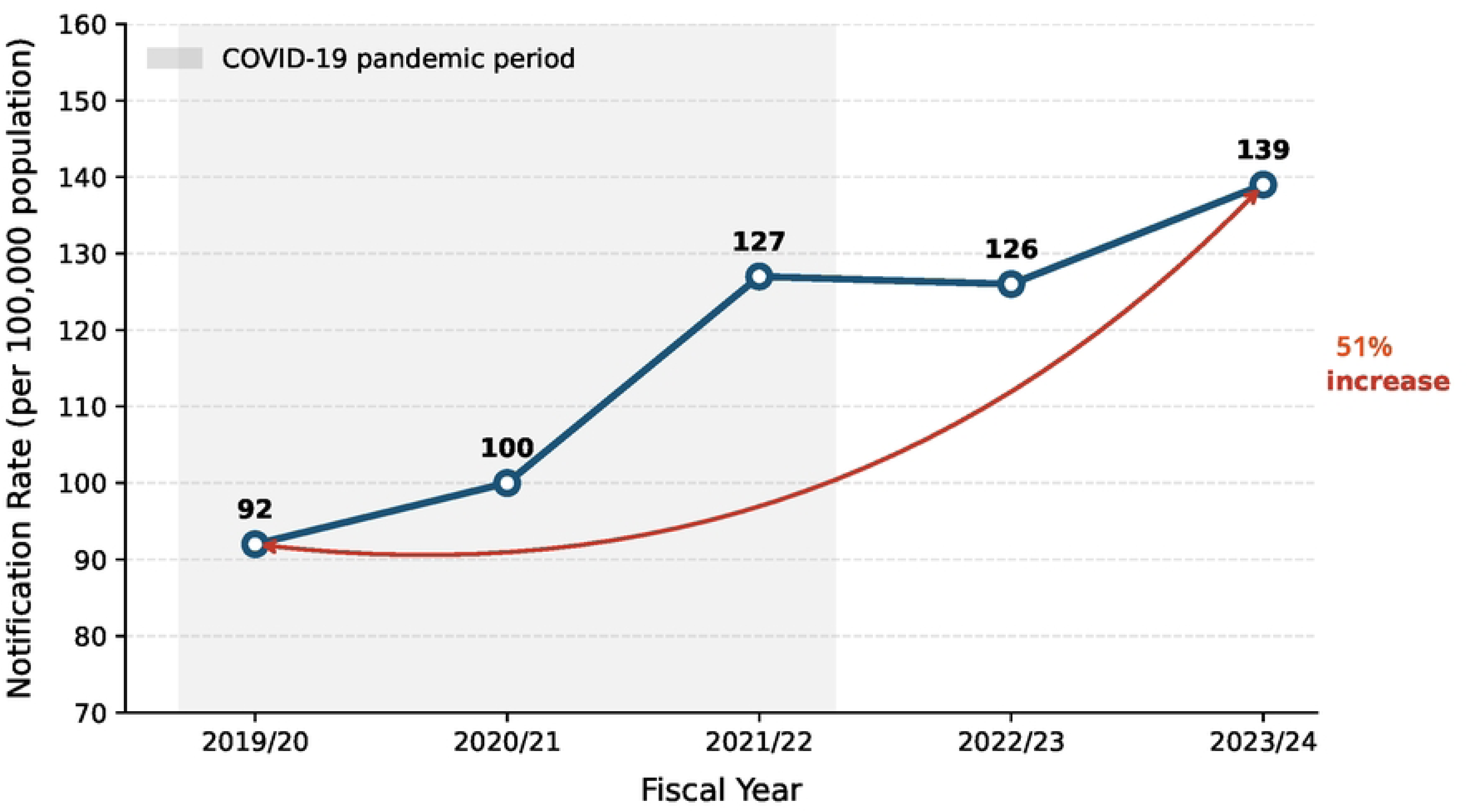
Temporal trends in tuberculosis notification rates, Nepal, fiscal years 2019/20-2023/24. The blue line shows annual case notification rates (per 100,000 population) across five fiscal years. Gray shading indicates the COVID-19 pandemic period (FY 2019/20-2021/22). The red arc illustrates the overall 51% increase from 92 to 139 per 100,000 between FY 2019/20 and FY 2023/24. Data source: National Tuberculosis Control Centre, Ministry of Health and Population, Nepal.

**Table 1.** Trends in Demographic and Clinical Characteristics of Tuberculosis Cases - Nepal, FY 2019/20–2023/24.

| Characteristic | 2019/20 n (%) | 2023/24 n (%) | Absolute Change (n) | Relative Change (%) |
| --- | --- | --- | --- | --- |
| **Total cases** | 27,542 | 40,776 | +13,234 | +48.1 |
| **Sex** |  |  |  |  |
| Female | 10,220 (37.1) | 15,797 (38.7) | +5,577 | +54.6 |
| Male | 17,322 (62.9) | 24,979 (61.3) | +7,657 | +44.2 |
| **Age group (years)** |  |  |  |  |
| 0–4 | 651 (2.4) | 2,007 (4.9) | +1,356 | +208.3 |
| 5–14 | 966 (3.6) | 1,561 (3.8) | +595 | +61.6 |
| 15–24 | 4,939 (18.5) | 6,200 (15.1) | +1,261 | +25.5 |
| 25–34 | 4,504 (16.8) | 5,707 (13.9) | +1,203 | +26.7 |
| 35–44 | 3,637 (13.6) | 4,812 (11.7) | +1,175 | +32.3 |
| 45–54 | 3,690 (13.8) | 5,316 (13.0) | +1,626 | +44.1 |
| 55–64 | 3,709 (13.8) | 5,907 (14.4) | +2,198 | +59.3 |
| ≥65 | 4,589 (17.1) | 9,310 (22.8) | +4,721 | +102.9 |
| **Type of tuberculosis** |  |  |  |  |
| Bacteriologically confirmed pulmonary (PBC) | 15,153 (55.0) | 23,426 (57.5) | +8,273 | +54.6 |
| Clinically diagnosed pulmonary (PCD) | 3,784 (13.7) | 6,237 (15.3) | +2,453 | +64.8 |
| Extrapulmonary (EP) | 8,605 (31.2) | 11,113 (27.3) | +2,508 | +29.1 |
| **Treatment history** |  |  |  |  |
| New | 25,116 (94.1) | 37,136 (90.9) | +12,020 | +47.9 |
| Relapse | 1,568 (5.8) | 3,034 (7.4) | +1,466 | +93.5 |
| Previously treated (other) | 1 (0.0) | 650 (1.5) | +649 | - |

### Space-Time Cluster Analysis

Retrospective space-time analysis identified four significant high-risk clusters with RR ≥1.5 (all p<0.001) (Table 2, Fig 2).

**Fig 2.**
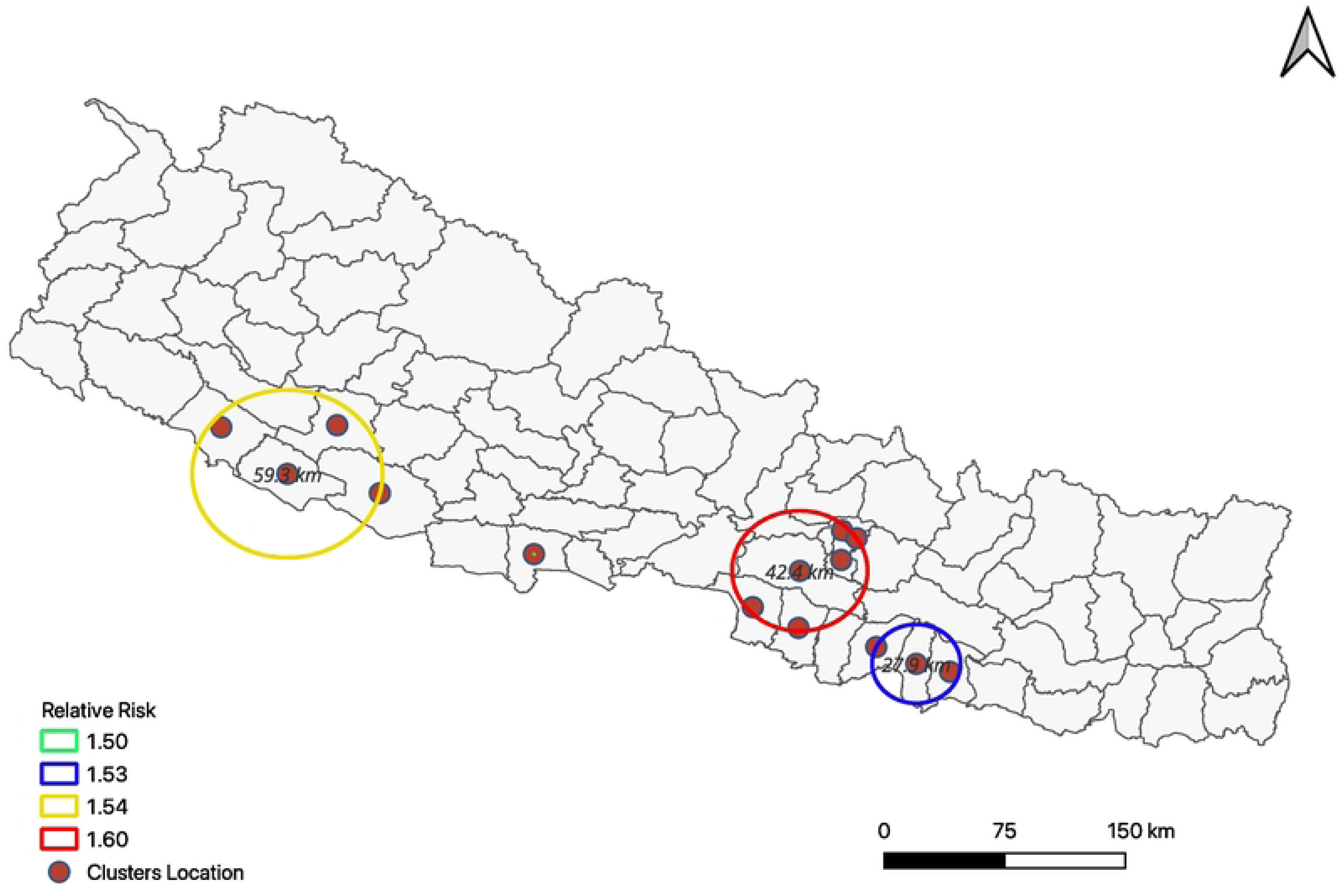
Geographic distribution of space-time tuberculosis clusters, Nepal, fiscal years 2019/20-2023/24. Map shows all 77 districts coloured by space-time cluster membership identified by SaTScan scan statistics (relative risk threshold ≥1.5, p<0.001). Cluster 1: Kathmandu Valley, 6 districts (dark red, RR=1.60, FY 2021/22–2023/24); Cluster 2: Western Terai and adjacent Hills, 4 districts (orange, RR=1.54, FY 2021/22–2023/24); Cluster 3: Eastern Terai, 3 districts (yellow, RR=1.53, FY 2022/23–2023/24); Cluster 4: Rupandehi (light yellow, RR=1.51, FY 2023/24). Non-cluster districts shown in gray.

**Table 2.** Space-Time Clusters of Tuberculosis Notifications Identified by Scan Statistics, Nepal, 2019/20–2023/24*.

| Cluster | Districts (n) | District Names (District Code) | Time Period (Fiscal Year) | Cases Observed | Cases Expected | RR | LLR | P Value |
| --- | --- | --- | --- | --- | --- | --- | --- | --- |
| 1 (MLC) | 6 | Kathmandu (306), Lalitpur (308), Bhaktapur (307), Makwanpur | FY 2021/22–2023/24 (Years 3–5) | 26,354 | 17,458 | 1.60 | 2218.1 | <0.001 |
|  |  | (312), Bara<br>(207), Parsa<br>(208) |  |  |  |  |  |  |
| 2 | 4 | Banke (511),<br>Salyan (609),<br>Bardiya (512),<br>Dang (510) | FY<br>2021/22-2023/24<br>(Years<br>3-5) | 10,433 | 6,917 | 1.54 | 809.3 | <0.001 |
| 3 | 3 | Dhanusha<br>(203),<br>Mahottari<br>(204), Sarlahi<br>(205) | FY<br>2022/23-2023/24<br>(Years<br>4-5) | 8,646 | 5,751 | 1.53 | 655.5 | <0.001 |
| 4 | 1 | Rupandehi<br>(508) | FY<br>2023/24<br>(Year 5) | 2,012 | 1,341 | 1.51 | 146.4 | <0.001 |

\*FY, fiscal year; LLR, log-likelihood ratio, MLC, most likely cluster; RR, relative risk. Scan parameters: maximum spatial cluster 20% of population, maximum temporal cluster 60% of study period (3 years), RR threshold $\geq 1.5$ , 999 Monte Carlo replications.

The most likely cluster comprised Kathmandu, Lalitpur, Bhaktapur, Makwanpur, Bara, and Parsa during FY 2021/22–FY 2023/24, with 26,354 observed versus 17,458 expected notifications (RR=1.60; LLR=2,218.1; p<0.001). These districts represented approximately 17% of Nepal’s population and accounted for 22.7% of all notifications during FY 2021/22–FY 2023/24. The cluster encompassed approximately 5.1 million people based on FY 2023/24 population estimates.

Three secondary clusters were also identified. Cluster 2 comprised four districts in western Nepal during FY 2021/22–FY 2023/24 (RR=1.54; LLR=809.3; p<0.001). Cluster 3 comprised three Eastern Terai districts during FY 2022/23–FY 2023/24 (RR=1.53; LLR=655.5; p<0.001). Cluster 4 comprised Rupandehi district during FY 2023/24 (RR=1.51; LLR=146.4; p<0.001). The detected clusters therefore varied in geographic extent and duration, with the primary and western clusters spanning three fiscal years, the Eastern Terai cluster spanning two years, and the Rupandehi cluster occurring during the final year.

### Purely Spatial Analysis

Purely spatial analysis, aggregating notifications across the five-year study period, identified five statistically significant clusters (Table 3, Fig 3).

**Fig 3.**
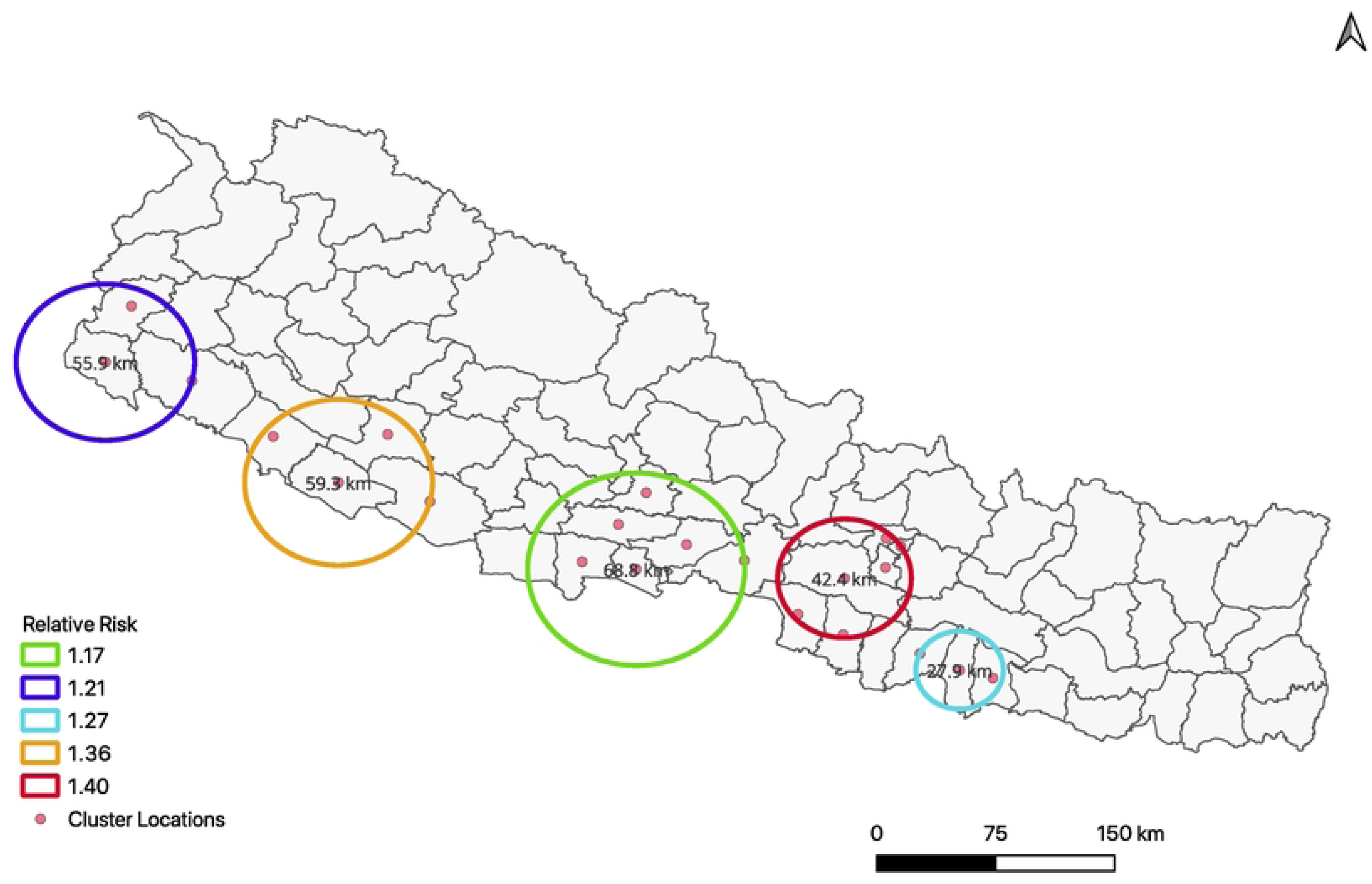
Compound epidemic pattern: integration of space-time, purely spatial, and purely temporal tuberculosis analyses, Nepal, 2019/20–2023/24. Panel A (left): Space-time cluster map showing four significant clusters (RR 1.51–1.60) concentrated in fiscal years 2021/22–2023/24. Panel B (centre): Purely spatial cluster map showing five persistent baseline elevation areas (RR 1.17–1.41) aggregated across all five fiscal years. Panel C (right): Conceptual diagram illustrating the compound epidemic relationship — baseline spatial inequality (RR=1.41) combined with national temporal surge (RR=1.26) produced concentrated space-time clusters (RR=1.60).

**Table 3.** Purely Spatial and Purely Temporal Tuberculosis Clusters, Nepal, 2019/20.

| Analysis Type | Geographic/Temporal Scope | District included | Cases Observed | Cases Expected | RR | LLR | P Value |
| --- | --- | --- | --- | --- | --- | --- | --- |
| **Purely Spatial (5-year aggregate)** |  |  |  |  |  |  |  |
| Cluster 1 | Kathmandu Valley region | Makawanpur, Lalitpur, Kathmandu, Parsa, Bara, Bhaktapur | 38,646 | 29,384 | 1.41 | 1634.0 | <0.001 |
| Cluster 2 | Western Terai and adjacent Hills | Banke, Salyan, Bardiya, Dang | 15,354 | 11,548 | 1.36 | 613.3 | <0.001 |
| Cluster 3 | Eastern Terai | Mahottari, Dhanusa, Sarlahi | 17,883 | 14,332 | 1.28 | 447.9 | <0.001 |
| Cluster 4 | Mid-Western region | Nawalparasi West, Palpa, Rupandehi, Nawalparasi East, Syangja, Chitawan | 20,698 | 17,999 | 1.17 | 216.8 | <0.001 |
| Cluster 5 | Far-Western | Kanchanpur, Dadeldhura, Kailali | 11,117 | 9,279 | 1.21 | 181.5 | <0.001 |
| **Purely Temporal (nationwide)** |  |  |  |  |  |  |  |
| Cluster 1 | All districts, Years 4-5 (2022/23–2023/24) | All 77 districts | 78,223 | 68,443 | 1.26 | 1144.0 | 0.001 |

\***LLR**, log-likelihood ratio; **RR**, relative risk

Three clusters geographically overlapped with areas identified in the space-time analysis. The largest cluster included Kathmandu, Lalitpur, Bhaktapur, Makawanpur, Bara, and Parsa, with an RR of 1.41 over the full study period. A second cluster comprised districts in western Nepal (RR=1.36), while a third included districts in the eastern Terai (RR=1.28). Two additional clusters were identified in the spatial-only analysis: a cluster comprising Nawalparasi East, Nawalparasi West, Palpa, Rupandehi, Syangja, and Chitawan (RR=1.17), and a Far-Western cluster comprising Kanchanpur, Dadeldhura, and Kailali (RR=1.21).

The spatial-only results therefore identified several areas with elevated notified TB burden over the aggregated five-year period, including areas that did not meet the RR≥1.5 reporting criterion in the primary space-time analysis. For the districts comprising the primary space-time cluster, the aggregated five-year spatial RR was 1.41 compared with an RR of 1.60 during FY 2021/22–FY 2023/24. This difference indicates that the elevated notification pattern in these districts was more pronounced during the later years of the study period.

### Purely Temporal Analysis (National Trend)

Purely temporal analysis identified one statistically significant high-notification period spanning FY 2022/23–FY 2023/24 (years 4–5), with 78,223 observed compared with 68,443 expected cases nationwide (RR=1.26, LLR=1,144.0, p=0.001) (Table 3). This indicates that national TB notifications were approximately 26% higher than expected during the final two fiscal years of the study period. No statistically significant high-or low-notification periods were identified for the remaining periods.

### Sensitivity Analyses

Sensitivity analyses showed that the principal space-time clustering pattern was broadly robust to alternative scanning-window specifications, although the geographic extent of clusters varied with the maximum spatial window. Increasing the maximum spatial window from 20% to 30% and 50% resulted in larger primary clusters while retaining substantial overlap with the core districts identified in the primary analysis. Changing the maximum temporal window from 60% to 50% also produced overlapping cluster patterns. An additional analysis using a 50% spatial and 50% temporal window without an RR reporting restriction identified additional lower-RR clusters, indicating that the primary RR ≥1.5 criterion primarily emphasized the more pronounced high-notification clusters. Detailed results of the alternative specifications are provided in Appendix Table 2.

## DISCUSSION

This nationwide spatiotemporal analysis identified three principal findings. First, national TB notification rates increased by 51%, from 92 to 139 per 100,000 population between FY 2019/20 and FY 2023/24, with four significant space-time clusters (RR 1.51–1.60) concentrated during FY 2021/22–FY 2023/24. Second, these clusters overlapped geographically with areas showing persistently elevated notified TB rates in the purely spatial analysis (RR 1.17–1.41), indicating substantial and persistent geographic heterogeneity in notified TB burden. Third, the purely temporal analysis identified a 26% increase in notifications during FY 2022/23–FY 2023/24, demonstrating a substantial nationwide increase during the later post-COVID recovery period. Together, these findings indicate that the national increase in TB notifications occurred alongside persistent geographic differences in notified TB burden. Similar geographic heterogeneity in TB notifications during the post-pandemic recovery period has been reported in Brazil, South Africa, and Indonesia [25–27]. By integrating space-time, purely spatial, and purely temporal scan statistics, this study characterizes both persistent geographic variation and temporal changes in TB notifications during Nepal’s post-COVID recovery period.

An important observation was that several districts identified as space-time clusters also showed elevated notified TB rates in the purely spatial analysis. This suggests that the increase in notifications during the post-COVID period occurred partly within areas that already experienced relatively high notified TB burden. From a programmatic perspective, areas with persistent elevation may require sustained TB control efforts in addition to intensified activities during periods of increased notifications. The Kathmandu-centered cluster, comprising Kathmandu, Lalitpur, Bhaktapur, Makawanpur, Bara, and Parsa districts, represented the most prominent space-time cluster identified in this study. These areas include major urban and peri-urban population centres characterized by substantial population mobility and socioeconomic heterogeneity [29,30], factors that may influence TB detection and notification patterns. However, the present ecological analysis did not directly evaluate the contribution of these factors to the observed clustering [15,31].

The Kathmandu-centered cluster emerged during FY 2021/22–FY 2023/24, following the period of major COVID-19-related disruption to healthcare access and TB services in Nepal [32]. Disruptions to diagnostic and treatment services during the pandemic may have delayed case detection and treatment initiation [33], while subsequent restoration of services and intensified case-finding activities may have increased detection of previously undiagnosed cases [34]. Similar mechanisms may have contributed to the increase in notifications observed in other high-notification areas. However, the present data cannot distinguish between increased underlying TB incidence and improved case detection. Therefore, the observed post-COVID increase should be interpreted as an increase in notified TB burden rather than direct evidence of increased TB incidence or transmission.

The temporal findings provide additional context for the geographic clusters. The significant nationwide increase in TB notifications during FY 2022/23–FY 2023/24 (RR=1.26) indicates that the post-COVID recovery involved a broad increase in notifications across Nepal, while the space-time analysis demonstrated substantial geographic heterogeneity, with selected districts experiencing higher notified burden than the national pattern (Tables 2 and 3). Purely spatial analysis further showed that geographic concentration persisted beyond the primary space-time clusters; for example, the Kathmandu Valley had a five-year RR of 1.41 compared with 1.60 during FY 2021/22–FY 2023/24, suggesting that the recent increase occurred in an area with an already elevated notified TB burden. Sensitivity analyses supported the robustness of the principal clustering pattern across alternative scanning-window specifications, although cluster boundaries varied with the maximum spatial window (Appendix Table 2). Analyses without the RR ≥1.5 reporting threshold additionally identified lower-RR clusters, indicating that the threshold emphasized more pronounced elevations rather than statistical significance. An important consideration when interpreting these findings is that TB notifications are not direct measures of TB incidence [35]. Changes in notification rates may reflect variations in healthcare utilization, diagnostic capacity, active case-finding activities, reporting completeness, and surveillance performance in addition to changes in underlying disease incidence [15]. Consequently, the identified clusters should be interpreted as areas of elevated notified TB burden rather than definitive indicators of increased TB incidence or transmission. In particular, the observed increase following the COVID-19 pandemic may reflect a combination of service recovery, improved case detection, delayed diagnosis, changes in healthcare-seeking behaviour, and underlying changes in disease occurrence. Distinguishing these mechanisms would require additional data beyond the scope of the present surveillance-based analysis.

### Strengths

This study has several strengths. First, it used national surveillance data covering all 77 districts of Nepal and the national population, providing comprehensive geographic coverage. Second, the five-year study period encompassed the pre-COVID period, COVID-19-related disruption, and subsequent recovery, allowing changes in TB notification patterns to be examined within a broader temporal context. Third, integration of space-time, purely spatial, and purely temporal scan statistics enabled assessment of persistent geographic variation alongside temporal changes in notifications. Fourth, sensitivity analyses using alternative spatial and temporal scanning parameters demonstrated broad robustness of the principal clustering pattern, although the geographic extent of some clusters varied according to the scanning window. Finally, the use of SaTScan version 10.3.2 and a prespecified analytical framework provides a reproducible approach that can be adapted to other settings with comparable surveillance data.

### Limitations

Several limitations should be considered. First, notification data reflect both disease occurrence and case detection; therefore, observed increases cannot be interpreted as direct measures of changes in TB incidence. Differences in healthcare access, diagnostic capacity, active case finding, reporting completeness, and surveillance performance may contribute to geographic variation in notifications. Second, the district-level ecological design cannot capture within-district heterogeneity, particularly in densely populated urban areas, and the findings should not be interpreted as individual-level associations. Finer-resolution analyses at the municipality or ward level may provide more precise information for local intervention planning. Third, the discrete Poisson model used in SaTScan does not explicitly account for potential spatial dependence or unmeasured heterogeneity in TB risk. Fourth, individual-level risk factors and clinical characteristics could not be incorporated because the analysis used aggregated surveillance data. Fifth, the RR ≥1.5 criterion was used as a prespecified reporting threshold to emphasize clusters with relatively large elevations in notified TB rates; it was not a criterion for statistical significance. Clusters with more modest relative risks may also have public health relevance and were observed under alternative analytical specifications. Finally, the observational ecological design does not permit causal attribution of the observed notification patterns to specific COVID-19-related disruptions, interventions, health-service changes, or policy measures.

### Public health implications

These findings support geographically targeted TB control alongside universal national services. The identified space-time clusters may benefit from intensified case finding, diagnostic capacity, contact investigation, and timely treatment, while persistently high-burden areas require sustained programmatic attention. The 26% increase in national notifications during FY 2022/23–FY 2023/24 also highlights the need to sustain post-COVID service recovery and strengthen routine spatial surveillance to guide resource allocation. The distribution of clusters in the western and eastern Terai further supports consideration of population mobility and cross-border coordination, although cross-border transmission was not directly assessed in this study [36].

## Conclusion

TB notifications increased substantially in Nepal between FY 2019/20 and FY 2023/24, with significant space-time clustering concentrated in specific geographic areas during the later COVID-19 recovery period. The coexistence of a nationwide temporal increase with persistent geographic concentration indicates that the post-COVID increase in notified TB burden was geographically heterogeneous. Combining space-time, purely spatial, and purely temporal surveillance analyses can help identify areas requiring sustained or intensified programmatic attention. However, because notification data reflect both disease occurrence and case detection, the identified clusters should not be interpreted as direct evidence of increased TB incidence or transmission. Strengthening routine surveillance, case detection, and geographically informed resource allocation may improve the responsiveness and efficiency of TB control efforts in Nepal.

## Supporting information

**Appendix Table 1.** Complete district-level tuberculosis data, cluster membership, and geographic information, Nepal, 2019/20–2023/24.

| **District Code (fid)** | **District Name** | **Province** | **Cluster Assignment** | **Cluster Time Period** | **Population (5-yr average)** | **Total Cases (5 years)** | **Average Annual CNR** | **RR in Cluster** | **Geographic Zone** |
| --- | --- | --- | --- | --- | --- | --- | --- | --- | --- |
| 101 | Taplejung | Koshi | None | — | 123,309 | 206 | 33.4 | — | Mountain |
| 102 | Sankhuwasabha | Koshi | None | — | 157,439 | 297 | 37.7 | — | Mountain |
| 103 | Solukhumbu | Koshi | None | — | 103,431 | 239 | 46.2 | — | Mountain |
| 104 | Okhaldhunga | Koshi | None | — | 144,001 | 451 | 62.6 | — | Hill |
| 105 | Khotang | Koshi | None | — | 172,516 | 197 | 22.8 | — | Hill |
| 106 | Bhojpur | Koshi | None | — | 155,680 | 297 | 38.2 | — | Hill |
| 107 | Dhankuta | Koshi | None | — | 159,041 | 316 | 39.7 | — | Hill |
| 108 | Terhathum | Koshi | None | — | 93,047 | 149 | 32.0 | — | Hill |
| 109 | Panchthar | Koshi | None | — | 181,933 | 236 | 26.0 | — | Hill |
| 110 | Ilam | Koshi | None | — | 293,000 | 685 | 46.8 | — | Hill |
| 111 | Jhapa | Koshi | None | — | 975,398 | 5,538 | 113.6 | — | Terai |
| 112 | Morang | Koshi | None | — | 1,134,374 | 5,892 | 103.9 | — | Terai |
| 113 | Sunsari | Koshi | None | — | 933,683 | 4,959 | 106.3 | — | Terai |
| 114 | Udayapur | Koshi | None | — | 349,112 | 1,439 | 82.4 | — | Hill |
| 201 | Saptari | Madhesh | None | — | 715,083 | 2,809 | 78.6 | — | Terai |
| 202 | Siraha | Madhesh | None | — | 733,796 | 3,563 | 97.2 | — | Terai |
| **203** | **Dhanusha** | **Madhesh** | **Cluster 3** | **FY**<br>**2022/23–**<br>**2023/24** | **864,982** | **6,529** | **151.0** | **1.53** | **Terai** |
| **204** | **Mahottari** | **Madhesh** | **Cluster 3** | **FY**<br>**2022/23–**<br>**2023/24** | **716,757** | **5,120** | **142.9** | **1.53** | **Terai** |
| **205** | **Sarlahi** | **Madhesh** | **Cluster 3** | **FY**<br>**2022/23–**<br>**2023/24** | **880,347** | **6,234** | **141.6** | **1.53** | **Terai** |
| 206 | Rautahat | Madhesh | None | — | 841,191 | 5,120 | 121.7 | — | Terai |
| **207** | **Bara** | **Madhesh** | **Cluster 1** | **FY**<br>**2021/22–**<br>**2023/24** | **789,812** | **6,193** | **156.9** | **1.60** | **Terai** |
| **208** | **Parsa** | **Madhesh** | **Cluster 1** | **FY**<br>**2021/22–**<br>**2023/24** | **683,800** | **5,064** | **148.2** | **1.60** | **Terai** |
| 301 | Dolakha | Bagmati | None | — | 177,682 | 484 | 54.5 | — | Hill |
| 302 | Sindhupalchok | Bagmati | None | — | 273,504 | 1,008 | 73.8 | — | Hill |
| 303 | Rasuwa | Bagmati | None | — | 45,833 | 164 | 71.6 | — | Hill |
| 304 | Dhading | Bagmati | None | — | 334,452 | 1,411 | 84.4 | — | Hill |
| 305 | Nuwakot | Bagmati | None | — | 271,865 | 995 | 73.3 | — | Hill |
| **306** | **Kathmandu** | **Bagmati** | **Cluster 1** | **FY**<br>**2021/22–**<br>**2023/24** | **2,138,922** | **17,075** | **159.6** | **1.60** | **Hill (Urban)** |
| **307** | **Bhaktapur** | **Bagmati** | **Cluster 1** | **FY**<br>**2021/22–**<br>**2023/24** | **415,109** | **2,921** | **140.7** | **1.60** | **Hill (Urban)** |
| **308** | **Lalitpur** | **Bagmati** | **Cluster 1** | **FY**<br>**2021/22–**<br>**2023/24** | **599,099** | **4,131** | **137.9** | **1.60** | **Hill (Urban)** |
| 309 | Kavrepalanchok | Bagmati | None | — | 380,742 | 1,546 | 81.2 | — | Hill |
| 310 | Ramechhap | Bagmati | None | — | 184,027 | 527 | 57.3 | — | Hill |
| 311 | Sindhuli | Bagmati | None | — | 304,785 | 1,543 | 101.3 | — | Hill |
| **312** | **Makwanpur** | **Bagmati** | **Cluster 1** | **FY**<br>**2021/22–**<br>**2023/24** | **432,700** | **3,262** | **150.9** | **1.60** | **Hill/Terai** |
| 313 | Chitwan | Bagmati | None | — | 718,409 | 5,062 | 141.0 | — | Terai |
| 401 | Gorkha | Gandaki | None | — | 247,865 | 1,028 | 83.0 | — | Hill |
| 402 | Manang | Gandaki | None | — | 6,000 | 7 | 23.3 | — | Mountain |
| 403 | Mustang | Gandaki | None | — | 13,586 | 42 | 61.8 | — | Mountain |
| 404 | Myagdi | Gandaki | None | — | 107,840 | 264 | 49.0 | — | Hill |
| 405 | Kaski | Gandaki | None | — | 600,183 | 2,999 | 99.9 | — | Hill |
| 406 | Lamjung | Gandaki | None | — | 159,985 | 608 | 76.1 | — | Hill |
| 407 | Tanahu | Gandaki | None | — | 332,882 | 1,412 | 84.9 | — | Hill |
| 408 | Nawalparasi<br>East | Gandaki | None | — | 371,573 | 2,536 | 136.6 | — | Terai |
| 409 | Syangja | Gandaki | None | — | 249,990 | 1,248 | 99.9 | — | Hill |
| 410 | Parbat | Gandaki | None | — | 137,111 | 343 | 50.1 | — | Hill |
| 411 | Baglung | Gandaki | None | — | 262,318 | 1,044 | 79.6 | — | Hill |
| 501 | Rukum East | Lumbini | None | — | 57,939 | 246 | 84.9 | — | Hill |
| 502 | Rolpa | Lumbini | None | — | 275,358 | 1,510 | 109.7 | — | Hill |
| 503 | Pyuthan | Lumbini | None | — | 235,481 | 1,696 | 144.2 | — | Hill |
| 504 | Gulmi | Lumbini | None | — | 247,916 | 1,191 | 96.2 | — | Hill |
| 505 | Arghakhanchi | Lumbini | None | — | 185,410 | 1,068 | 115.3 | — | Hill |
| 506 | Palpa | Lumbini | None | — | 244,332 | 1,685 | 137.9 | — | Hill |
| 507 | Nawalparasi West | Lumbini | None | — | 384,573 | 2,303 | 119.8 | — | Terai |
| **508** | **Rupandehi** | **Lumbini** | **Cluster 4** | **FY 2023/24** | **1,111,540** | **7,864** | **141.5** | **1.51** | **Terai** |
| 509 | Kapilbastu | Lumbini | None | — | 685,962 | 4,010 | 117.0 | — | Terai |
| **510** | **Dang** | **Lumbini** | **Cluster 2** | **FY 2021/22–2023/24** | **669,448** | **5,978** | **178.6** | **1.54** | **Terai** |
| **511** | **Banke** | **Lumbini** | **Cluster 2** | **FY 2021/22–2023/24** | **611,724** | **5,051** | **165.1** | **1.54** | **Terai** |
| **512** | **Bardiya** | **Lumbini** | **Cluster 2** | **FY 2021/22–2023/24** | **470,342** | **3,316** | **141.0** | **1.54** | **Terai** |
| 601 | Dolpa | Karnali | None | — | 43,167 | 166 | 77.0 | — | Mountain |
| 602 | Mugu | Karnali | None | — | 65,982 | 223 | 67.6 | — | Mountain |
| 603 | Humla | Karnali | None | — | 57,440 | 118 | 41.1 | — | Mountain |
| 604 | Jumla | Karnali | None | — | 122,187 | 467 | 76.5 | — | Mountain |
| 605 | Kalikot | Karnali | None | — | 152,019 | 445 | 58.5 | — | Hill |
| 606 | Dailekh | Karnali | None | — | 271,223 | 792 | 58.4 | — | Hill |
| 607 | Jajarkot | Karnali | None | — | 194,719 | 570 | 58.6 | — | Hill |
| 608 | Rukum West | Karnali | None | — | 148,495 | 806 | 108.6 | — | Hill |
| **609** | **Salyan** | **Karnali** | **Cluster 2** | **FY**<br>**2021/22–**<br>**2023/24** | **234,833** | **1,009** | **86.0** | **1.54** | **Hill** |
| 610 | Surkhet | Karnali | None | — | 420,677 | 2,627 | 124.9 | — | Hill/Terai |
| 701 | Bajura | Sudurpashchim | None | — | 145,974 | 681 | 93.3 | — | Hill |
| 702 | Bajhang | Sudurpashchim | None | — | 202,093 | 781 | 77.3 | — | Hill |
| 703 | Darchula | Sudurpashchim | None | — | 138,651 | 634 | 91.5 | — | Hill |
| 704 | Baitadi | Sudurpashchim | None | — | 252,800 | 901 | 71.3 | — | Hill |
| 705 | Dadeldhura | Sudurpashchim | None | — | 147,472 | 638 | 86.6 | — | Hill |
| 706 | Doti | Sudurpashchim | None | — | 208,867 | 904 | 86.6 | — | Hill |
| 707 | Achham | Sudurpashchim | None | — | 251,929 | 803 | 63.8 | — | Hill |
| 708 | Kailali | Sudurpashchim | None | — | 930,553 | 6,446 | 138.5 | — | Terai |
| 709 | Kanchanpur | Sudurpashchim | None | — | 524,500 | 4,033 | 153.8 | — | Terai |

\***CNR**, case notification rate (per 100,000 population per year, averaged over 5 fiscal years); **FY**, fiscal year; **RR**, relative risk. Average annual CNR calculated as: (Total cases over 5 years / Total person-years) × 100,000. RR values shown only for districts within identified space-time clusters; represents RR of the cluster to which the district belongs. Geographic zones: Mountain (>3,000 m), Hill (600–3,000 m), Terai (<600 m). Urban classification indicates major metropolitan areas.

**Appendix Table 2.** Sensitivity analysis results SaTScan purely spatial and purely temporal analyses under alternative parameter specifications, Nepal, FY 2019/20–2023/24.

| **Analysis** | **Spatial Window** | **Temporal Window** | **Study Period** | **Clusters Detected (n)** | **Primary Cluster Districts** | **Primary Cluster Time Period** | **Primary Cluster RR** | **Key Observation** |
| --- | --- | --- | --- | --- | --- | --- | --- | --- |
| **PRIMARY** | **20%** | **60%** | **Years 1–5** | **4** | **6 (Kathmandu Valley region)** | **FY 2021/22–2023/24 (Years 3–5)** | **1.60** | **Reference analysis; 4 distinct geographically and temporally defined clusters** |
| Sensitivity 1 | 20% | **50%** | Years 1–5 | 4 | 6 (Kathmandu Valley region) | FY 2022/23–2023/24 (Years 4–5) | 1.58 | Same 4 geographic areas detected; Kathmandu cluster onset truncated to Year 4, missing Year 3 emergence; Western cluster truncated to Years 3–4, missing Year 5; confirms 60% temporal window needed to capture full surge arc |
| Sensitivity 2 | **30%** | 60% | Years 1–5 | 3 | **11 (Kathmandu Valley + Eastern Terai merged)** | FY 2021/22–2023/24 (Years 3–5) | 1.55 | Larger window merges epidemiologically distinct Kathmandu Valley and Eastern Terai clusters into single 82-km radius cluster; loses actionable geographic distinction; validates 20% spatial specification |
| Sensitivity 3 | **50%** | 60% | Years 1–5 | 3 | **12 (Very large merged cluster)** | FY 2021/22–2023/24 (Years 3–5) | 1.57 | Very large cluster (81 km radius, 9.0 million population, 31% of Nepal) spans multiple ecological zones; not epidemiologically meaningful for targeted intervention; further validates 20% spatial specification |
| Sensitivity 4 (Pandemic exclusion) | 20% | 60% | **Years 3–5 only** | 3 | 1 per cluster (single districts) | Year 5 / Year 3 / Year 5 | 1.59 / 1.63 / 1.53 | Restricting to post-pandemic years detects same geographic areas (Bara/207, Dang/510, Dhanusha/203) but as individual district-year observations rather than multi-district clusters; confirms underlying geographic signals are robust and present in post-pandemic period alone, while demonstrating that multi-district cluster detection requires full 5-year period for adequate baseline comparison |

**\*All analyses: Discrete Poisson model, $RR \geq 1.5$ threshold, 999 Monte Carlo replications, circular window, no geographic overlap for secondary clusters.**
**Primary analysis specified a priori; all sensitivity analyses conducted post-hoc to assess robustness.**
**RR, relative risk; FY, fiscal year.**

**Appendix Table 3.** Time period coding, fiscal year equivalents, and key contextual events, Nepal TB study.

| **Analysis Code (Generic Time)** | **Nepal Fiscal Year** | **Calendar Period** | **Start Date** | **End Date** |
| --- | --- | --- | --- | --- |
| **1** | 2019/20 | July 2019-July 2020 | July 16, 2019 | July 15, 2020 |
| **2** | 2020/21 | July 2020-July 2021 | July 16, 2020 | July 15, 2021 |
| **3** | 2021/22 | July 2021-July 2022 | July 16, 2021 | July 15, 2022 |
| **4** | 2022/23 | July 2022-July 2023 | July 16, 2022 | July 15, 2023 |
| **5** | 2023/24 | July 2023-July 2024 | July 16, 2023 | July 15, 2024 |

\*CNR, case notification rate; TB, tuberculosis.
Calendar period approximations for international readability. Nepal's fiscal year begins mid-July (Shrawan 1 in Nepali calendar) and ends mid-July of the following calendar year, approximately 6 months offset from January-December calendar years. Key events compiled from Nepal Ministry of Health and Population COVID-19 response reports, National Tuberculosis Control Centre annual reports, and published literature on TB service disruption in Nepal during the pandemic period.

## ACKNOWLEDGEMENTS

We thank the National Tuberculosis Control Centre, Ministry of Health and Population, Government of Nepal, for providing access to TB surveillance data. We acknowledge the Central Bureau of Statistics, Nepal, for population data, and the district and provincial health offices for their continuous efforts in TB case detection and reporting.

## AUTHOR CONTRIBUTIONS

Conceptualization: Nabin Bisht, Rupam Bhatt, Pratik Rijal, Chiranjivi Adhikari

Data curation: Nabin Bisht, Harikishor Yadav, Samir Singh, Dilip Kumar Kalwar

Formal analysis: Nabin Bisht, Chiranjivi Adhikari

Funding acquisition: Not applicable

Investigation: Nabin Bisht, Rupam Bhatt, Pratik Rijal, Harikishor Yadav, Samir Singh, Dilip Kumar Kalwar, Rekha Bisht

Methodology: Nabin Bisht, Ravi Kumar Gupta, Chiranjivi Adhikari

Project administration: Nabin Bisht, Chiranjivi Adhikari

Resources: Chiranjivi Adhikari, Rekha Bisht

Software: Nabin Bisht, Ravi Kumar Gupta

Supervision: Chiranjivi Adhikari, Ravi Kumar Gupta

Validation: Nabin Bisht, Rupam Bhatt, Pratik Rijal, Rekha Bisht, Harikishor Yadav, Samir Singh, Dilip Kumar Kalwar, Chiranjivi Adhikari

Writing : original draft: Nabin Bisht

Writing, review & editing: Nabin Bisht, Rupam Bhatt, Pratik Rijal, Ravi Kumar Gupta, Rekha Bisht, Harikishor Yadav, Samir Singh, Dilip Kumar Kalwar, Chiranjivi Adhikari

## Data availability statement

The tuberculosis notification data analysed in this study were obtained from the National Tuberculosis Control Centre (NTCC), Ministry of Health and Population, Government of Nepal. These data are not publicly available due to institutional restrictions and data-sharing agreements. However, the datasets may be made available from the corresponding author upon reasonable request and with prior permission from the National Tuberculosis Control Centre. Sociodemographic data from the 2021 National Population and Housing Census are publicly available from the Central Bureau of Statistics, Nepal at https://nsonepal.gov.np/. SaTScan software is freely available at https://www.satscan.org/. Administrative boundary data are available from the National Geoportal at https://nationalgeoportal.gov.np/#/

